# Prevalence of Non-communicable diseases among the pregnant women in selected three teagardens of Sreemongol Upazila in Moulvibazar district: A Cross Sectional Study

**DOI:** 10.64898/2026.03.22.26348744

**Authors:** Abu Sayeed Md. Abdullah, Farjana Haq, Koustuv Dalal

## Abstract

**Background:** Bangladesh is experiencing emerging burden of Non-Communicable Diseases (NCDs). Non-communicable diseases (NCDs) are the emerging as major cause of morbidity and mortality, accounting for 61% of deaths in Bangladesh.

**Objective:** The study aims to describe the prevalence of NCDs among pregnant women in teagardens in Moulvibazar district.

**Methods:** Three teagardens of Sreemongol upazila in Moulvibazar district was selected randomly. The pregnant women were considered for collecting the NCD related information. A sample size of 86 was purposively selected based on relevant literature review. Data was collected by conducting face to face interview with the respondents through pre-tested semi-structured questionnaire. Data was analyzed with the help of SPSS Version 24 Software.

**Results:** For effective use of limited resources, an increased understanding of the shifting burden and better characterization of risk factors of NCDs including Hypertension is needed. Average age of the women attended for screening test was 23 (15-45) years. More than 47% women were found with Gravida 1. The mean duration of pregnancy was found 18.8 weeks. Above 24% percent of GDM women were found at low blood pressure but 2% were identified at high blood pressure. 28% were found underweight with BMI calculation but 11% were identified with overweight. The challenges tests for blood sugar findings of women were found 12.7% GDM positive (7.8-<11 mmol/L). About 16.5% had complications during pregnancy including anaemia, eclampsia, edema, diarrhoea etc.

**Conclusion:** A community based NCDs surveillance model could be developed through participation Government health managers, experts and stakeholders, which were taken by local health system for implementation.

## Introduction

Non-communicable diseases (NCDs) are those diseases that cannot be passed from person to person include cardiovascular diseases e.g. heart attacks and stroke, cancers, chronic respiratory diseases such as chronic obstructive pulmonary disease and asthma and diabetes **[1]**. Smoking, lack of physical activity, misuse of alcohol and unhealthy diets all increase the risk of dying from an NCD. Four key metabolic/physiological changes increase the risk of NCDs: raised blood pressure (hypertension), overweight/obesity, high blood glucose levels (hyperglycemia) and high levels of fat in the blood/hyperlipidemia **[2]**. In 2014, more than 1.9 billion adults were overweight; of these, over 600 million were obese **[3]**. About 39% of adults were overweight, and 13% were obese. 42 million children under five years of age were overweight or obese in 2013 **[4]**.

In 2013 a study found that 21.2% pregnant women were reported as obese with pregnancy BMI of >30 kg/m2. Above 40% and 32.8% pregnant women were reported as overweight and healthy with pregnancy BMI of 25–29.9 kg/m2 and 18.5–24.9 kg/m2, respectively **[5]**.

Diabetes is the leading cause of death and disability worldwide. The current prevalence of diabetes in Bangladesh is 5.5% and the expected rise to 8.2% by 2030. Gestational diabetes affects 3-10% of pregnancies. Gestational Diabetes Mellitus (GDM) is a very common disease occurs during pregnancy and has detrimental effect on both the mother and the baby **[6]**. Diabetes in pregnancy is a neglected cause of maternal mortality, where one in every six pregnancies was affected with GDM globally. About 88% GDM were found in low- and middle-income countries. One in every seven pregnant women suffers from hyperglycemia globally where 85% are GDM **[7]**. About 30% pregnant women may be affected but many of them are undiagnosed with the consequence of maternal and neonatal deaths. The current prevalence of 5.5% and the expected rise to 8.2% by 2030 renders diabetes in Bangladesh a major public health problem which has to be faced by the national health system in the near future **[8]**. Diabetes is associated with transitions in lifestyle and industrialization is more common in urban areas, but extends increasingly to rural settings. Of all outpatient visits, diabetes related consultations accounted for 23% of all outpatient patients in district hospitals, 25% at Upazila Health Complexes and alone for 31% of visits at primary care level in 2009 **[6]**.

In the past five years, there was an increased focus in Bangladesh on improving service delivery at primary level and referral centres including Union health and Family Welfare Centres as well as on community level through community clinics and community health workers **[9]**. The country was on track to reach its MDGs and has reduced its maternal mortality to a ratio of 194 **[10]**. Nevertheless, most programmes still focus on the provision of EmONC (Emergency Obstetric and Newborn Care) with poor focus on other maternal and newborn health related areas that are on the incline such as the impact of NCDs on MNH **[11]**. A new operational plan provides a structure for improving care with regards to preconception, pregnancy, childbirth and the immediate postpartum period. Areas prioritized for providing quality maternal and neonatal health services include populations with a high maternal mortality ratio, and areas of poverty, geographical and social disadvantages **[12]**. In Bangladesh there is a policy on non-communicable diseases and a national guideline on Maternal and Neonatal Health Strategies and Standard Operating Procedures has been developed for NCD management from community to district and division health care level **[11]**.

NCDs are emerging in Bangladesh as major cause of morbidity and mortality like many other developing countries **[13]**. It was estimated about 61% of deaths occurred due to NCDs in Bangladesh. The most common NCDs include cardiovascular diseases, diabetes mellitus, cancer, and chronic respiratory diseases **[14]**. A study in Bangladesh provides evidence of increasing trends in obesity among pregnant women, which poses possible health risks both for mother and child **[15]**. In the year 2015 a hospital-based study in Dhaka, Bangladesh, documented an overall incidence of hypertensive disorders of pregnancy to be 13.9% **[16]**. Bangladesh is one of the top ten contributors of diabetes cases globally and the numbers will double in the next 20 years. It is estimated that alone in Bangladesh more than two million people suffer from diabetes but are yet undiagnosed **[17]**. The prevalence of GDM ranges from 8.2% in rural Bangladesh to 12.9% but estimates depend on the diagnostic criteria used. Undiagnosed cases of women with diabetes in pregnancy are a key concern in Bangladesh, where the social status of women is low resulting in inequalities regarding healthcare access and nutrition **[18]**. Women often do not receive treatment for sickness, more often resort to self-medication, or seek care from unqualified practitioners **[19]**. The mother is at increased risk of developing obstetric complications like prolonged labour, prone to develop type-2 diabetes in future and the baby is born with overweight, cause of childhood obesity and later life development of type-2 diabetes **[20]**. It is estimated that alone in Bangladesh more than two million people suffer from diabetes but are yet undiagnosed. The existing community network can be used to refer all pregnancies identified at community level for antenatal care booking and diagnosis of NCDs to the facilities. The aim of the study is to access the prevalence of non-communicable diseases among the pregnant women in selected three teagardens in Moulvibazar district

## Methods

### Study Design

A facility based cross sectional study method was used to conduct the study.

### Study Setting

The study was conducted in three randomly selected teagardens in Sreemongol Upazila, Moulvibazar district, Bangladesh, between July and December 2019.

### Participants

Pregnant women attending antenatal care services in selected teagardens were included. Women who were severely ill or unwilling to participate were excluded.

### Sample Size

A total of 86 participants were included based on feasibility and prior similar studies.

Total 3 teagardens were selected randomly out of 45 teagardens in Sreemongol upazila of Moulvibazar district. Among them two were situated in Sathgaon union and one in Rajghat union. Two teagardens were with the ownership of Finley and one was within Sathgaon tea state [Table 1].

**Table 1:** Distribution of the samples according to the teagardens.

| *Upazila* | *Union* | *Ownership* | *Name of the Teagarden* | *No of pregnant women (Sample)* |
| --- | --- | --- | --- | --- |
| Sreemongol | Sathgaon | Sathgaon tea state | Sathgaon | 29 |
|  |  | Finley | Amrailchara | 32 |
|  | Rajghat | Finley | Rajghat | 25 |
| **Total** | **2** | **2** | **3** | **86** |

### Variables and Measurements

Primary outcomes included:

- Gestational diabetes mellitus (GDM)
- Hypertension
- Body mass index (BMI)

GDM was assessed using a single-step 75g OGTT with cut-offs of 7.8–11.0 mmol/L. Blood pressure and anthropometric measurements were obtained using standard procedures.

### Data Collection

Data were collected using a structured questionnaire, clinical assessments, and ANC record reviews. A structured pre-tested questionnaire was used to collect the data at the teagardens facilities by interviewing the respondent face to face to get the depth information from the hospital of selected 3 teagardens. Pregnant women came to the teagardens hospital to receive the ANC. The midwife of the teagarden hospital was assigned to provide the ANC with the necessary instruments. The data collector collected the information from interviewing the pregnant women at facility and observing and checking the ANC records at the facility. The standard protocol was followed to collect the information from women. The protocol maintained for detection of hypertension, obesity, diabetes related NCDs among pregnant women. The draft protocol for GDM screening program was presented and agreed at national advocacy meeting.

Single step OGTT i.e. blood sugar level 2hr after 75 gm glucose was performed for screening of GDM in pregnant women and the cut off value for this test is 7.8-11.1 mmol/L to detect GDM.

### Bias

Selection bias may exist due to facility-based purposive sampling. Measurement bias was minimized using standardized protocols.

### Statistical Analysis

A tabulation plan was prepared for quantitative data containing dummy tables as per the study objectives. SPSS data analysis programme was used to produce different uni-variate and bi-variate tables to address the study objectives. Descriptive analysis of different viable were also made using IBM SPSS version 22.0 for windows statistical software. Analyses were presented in the form of tables.

### Ethical Considerations

Ethical approval was obtained from the Ethical Review Committee of CIPRB [Memo: CIPRB/ERC/2016/010]. The study and tools were reviewed and finally approved by the honorable Supervisor of the researcher. Informed consent was taken from each of the response before the interview process. Participation of the respondent was voluntary and freedom was given to the respondent to response questions.

## Results

### Participant Characteristics

During the project period 86 pregnant women were considered for data collection from selected three gardens. The basic information includes the age, education, gravida, LMP, height, weight and pregnancy duration were collected. The hypertension related information; history of previous GDM, obesity related information was performed.

Average age of the women attended for screening test was 23 (15-45) years. Age of the women were mostly found with the age range of 20-24 years, the percentage of which is about 40%, whereas 25% women were found below 20 years. 23.5% women were at 25-29 years and 11.6% women were found above 30 years. Above 55% women were found below primary level of education. Only 8.7% women were found completed the secondary education. More than 47% of the pregnant women were found with Gravida 1, where about 37% with Gravida 2 and 16% women were found with Gravida 3 and above. The mean duration of pregnancy was found 18.8 weeks among the women participated in screening. Among them only 28% women were come between 24-28 weeks. Above 20% women were found below 12 weeks and about 50% women were found at 13-<24 weeks of pregnancy. 1.5% women were found above 28 weeks of pregnancy **[Table 2]**.

**Table 2:**
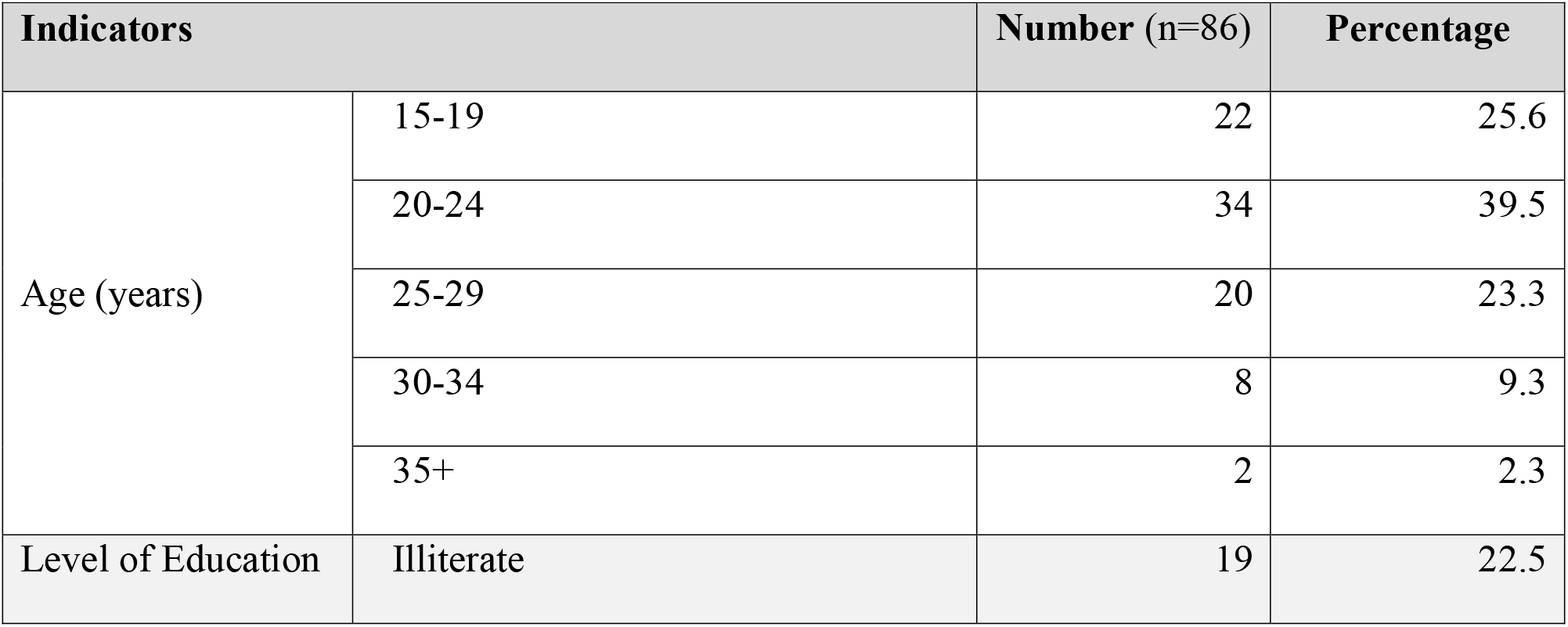
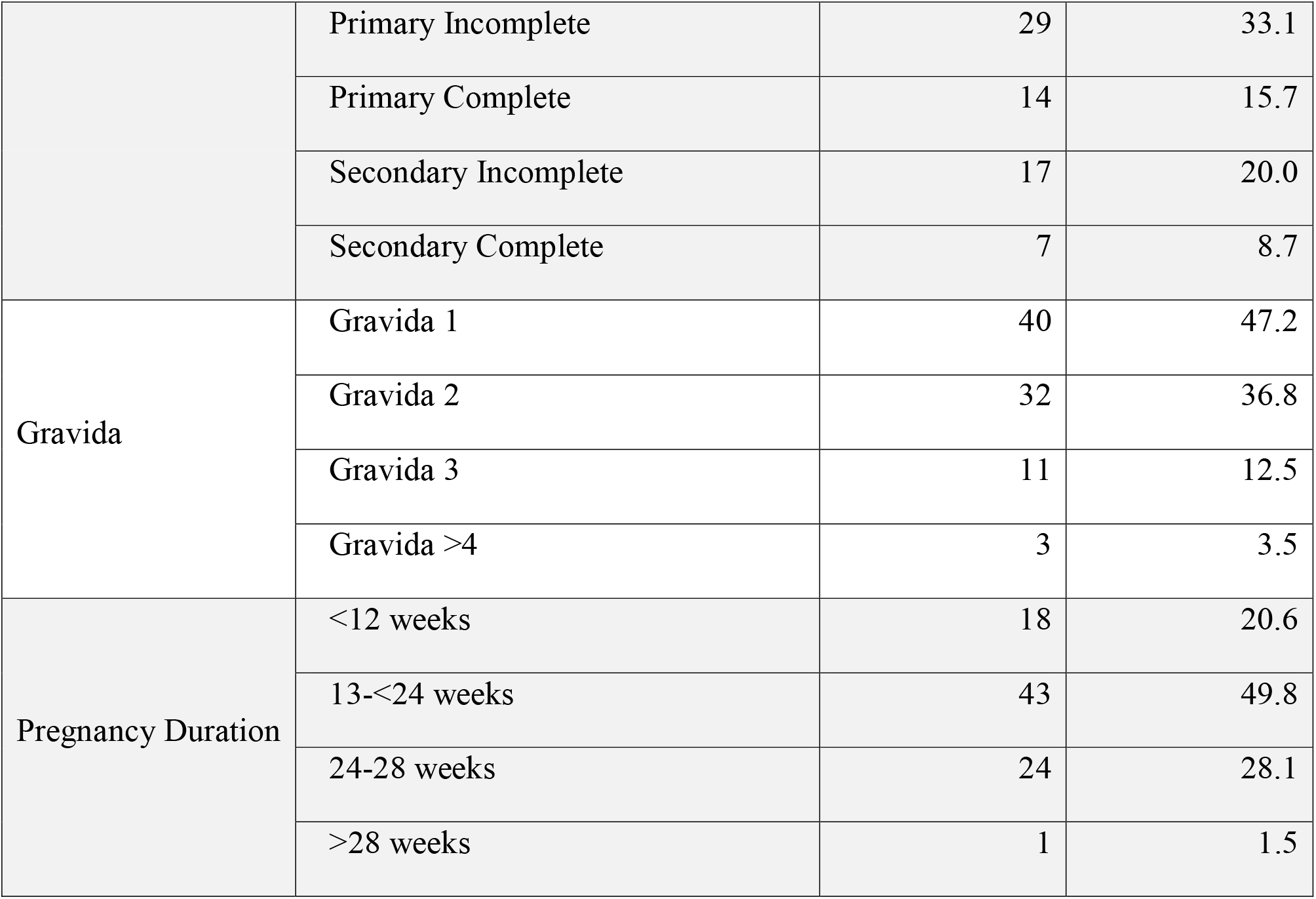
Distribution of women according to the Sociodemographic and Obstetric Characteristics.

| Indicators |  | Number (n=86) | Percentage |
| --- | --- | --- | --- |
| Age (years) | 15-19 | 22 | 25.6 |
|  | 20-24 | 34 | 39.5 |
|  | 25-29 | 20 | 23.3 |
|  | 30-34 | 8 | 9.3 |
|  | 35+ | 2 | 2.3 |
| Level of Education | Illiterate | 19 | 22.5 |
|  | Primary Incomplete | 29 | 33.1 |
|  | Primary Complete | 14 | 15.7 |
|  | Secondary Incomplete | 17 | 20.0 |
|  | Secondary Complete | 7 | 8.7 |
| Gravida | Gravida 1 | 40 | 47.2 |
|  | Gravida 2 | 32 | 36.8 |
|  | Gravida 3 | 11 | 12.5 |
|  | Gravida >4 | 3 | 3.5 |
| Pregnancy Duration | <12 weeks | 18 | 20.6 |
|  | 13-<24 weeks | 43 | 49.8 |
|  | 24-28 weeks | 24 | 28.1 |
|  | >28 weeks | 1 | 1.5 |

### Prevalence of NCDs

Blood pressure was measured among 86 women. Above twenty four percent of GDM women were found at low blood pressure. Although 74% women were found at normal blood pressure but 2% were identified at high blood pressure. Total 86 women had undergone 75 gm glucose challenges test. Based on the challenge tests for blood sugar, 73 women i.e 12.7% were found GDM positive (7.8 - <11 mmol/L). About 2% women were identified with pre-existing diabetes (≥11.1 mmol/L). About 67% women said they don’t know about NCD and NCD related complications for mother and baby, whereas 16.7% said there is no complication of NCDs and rest 16.7% said there are some complications of NCDs which effects mother and baby. Some of the respondent said new born can become abnormal, pregnant mothers may caught diabetes **[Table 3]**.

**Table 3:** Distribution of women according to the Blood Pressure, BMI, Blood Sugar, and NCD Status.

| Indicators |  | Number<br>(n=86) | Percentage |
| --- | --- | --- | --- |
| Blood Pressure | Normal BP (120/80) | 63 | 73.8 |
|  | Low BP (90/60) | 21 | 24.5 |
|  | High BP (140/90) | 2 | 1.7 |
| BMI findings | Underweight (below 18.5) | 24 | 28.0 |
|  | Healthy Weight (18.5-24.9) | 52 | 60.9 |
|  | Overweight (25-29.9) | 10 | 11.1 |
| Challenge test for blood sugar | Negative (<7.8 mmol/L) | 73 | 85.4 |
|  | GDM (7.8 - <11 mmol/L) | 11 | 12.7 |
|  | Pre-existing Diabetes (>11 mmol/L) | 2 | 1.9 |
| Knowledge on NCD | No | 58 | 66.6 |
|  | Yes | 28 | 33.4 |
| Complication for NCD among the known women | No | 14 | 16.7 |
|  | Yes | 14 | 16.7 |

#### Nutritional Status

Out of total women 28% were found underweight with BMI calculation. Though 61% women were at healthy weigh, but 11% were identified with overweight **[Table 3]**.

### NCDs related observation findings at facilities

Among 86 ANC observations in the facilities only 31.3% cases were found where blood sugar test related information was collected and 12.5% cases were identified with possible high level of blood sugar. Only 4.2% cases of ANC observation were found with receiving the NCDs related information where none of the cases found that the ANC provider counsel with the pregnant women about the NCD related risk factors. It was also observed that only 31.3% cases were asked either diabetes in her family members. Only 41.7% and 39.6% ANC were found where weight and height respectfully were measured from pregnant women. Moreover 29.2% and 12.5% cases of ANC were found where previous history of abortion and either last delivered baby’s weight more than 04 kg were taken respectively. No ANC were found where receiving the history of previous GDM. Among the total number of ANC 45.8% cases were found to measure the blood pressure of pregnant women. Only 2.1% and 12.5% cases were found to receive last delivery complication and either suffered from hypertension/Pre-eclampsia/PPH respectively. Only 2.1% cases were found with counsel on neonatal complication. No ANC were found where the provider counsel on NCD related information include future development of diabetes in life, disappear of NCD after pregnancy, period of pregnancy to diagnose NCDs. Even no ANC were found where capillary sugar test or urine sugar test or refer women to a laboratory for a sugar test. Diet plan was shared among the 50% cases of ANC and 29.2% cases were found where danger sign related leaflet was used for counseling. More than 58% cases of ANC were found where the provider mentioned the next appointment of receiving ANC **[Table 4]**.

**Table 4:** Distribution of NCD related observation findings among the pregnant women at facilities.

| Indicators | Number (n=86) | Percentage |
| --- | --- | --- |
| Information about sugar test in pregnancy | 27 | 31.3 |
| Either mentions the possibility of having high sugar levels only in pregnancy | 11 | 12.5 |
| Either speak about NCD | 4 | 4.2 |
| Either explore about risk factors for NCDs |  | - |
| Either receive weight | 36 | 41.7 |
| Either receive height (BMI) | 34 | 39.6 |
| Either ask for diabetes in family | 27 | 31.3 |
| Either ask if previous abortion | 25 | 29.2 |
| Either ask if previous baby more than 4 kg | 11 | 12.5 |
| Either check blood pressure | 39 | 45.8 |
| Either mentions potential complications of NCDs | 22 | 25 |
| Either counsel on delivery complications in general | 2 | 2.1 |
| Either counsel on Shoulder dystocia |  | - |
| Either counsel on Postpartum bleeding |  | - |
| Either counsel on Hypertension/ Preeclampsia | 3 | 3.1 |
| Either counsel on complications for the newborn | 2 | 2.1 |
| Either counsel on Macrosomia | 5 | 6.3 |
| Either counsel on Respiratory problems | 13 | 14.6 |
| Either counsel on NCDs related factors |  | - |
| Either mention that NCDs can be treated | 11 | 12.5 |
| Either show Diet chart | 43 | 50.0 |
| Either counsel on Insulin | 22 | 25.1 |
| Either provide leaflet for danger signs in pregnancy | 25 | 29.2 |
| Either give next appointment | 50 | 58.3 |

## Discussions

This study assessed the prevalence of selected non-communicable diseases (NCDs) among pregnant women in teagarden communities of Sreemongol, Bangladesh, and identified important gaps in antenatal care (ANC) services. The findings reveal a notable burden of gestational diabetes mellitus (GDM) (12.7%), along with smaller proportions of hypertension (1.7%) and overweight (11.1%), highlighting the emerging importance of NCDs in maternal health even in marginalized populations.

The prevalence of GDM observed in this study is consistent with findings from other studies conducted in Bangladesh, where estimates range from 8.2% in rural areas to approximately 12.9% depending on diagnostic criteria [21,22]. Globally, GDM affects approximately 3–10% of pregnancies, but higher rates are increasingly reported in low- and middle-income countries due to rapid epidemiological transitions [23]. The relatively high prevalence observed in this teagarden population suggests that even traditionally underserved and rural populations are not immune to the rising burden of metabolic disorders.

The low prevalence of hypertension (1.7%) in this study is lower than previously reported estimates in Bangladesh, where hypertensive disorders of pregnancy have been reported around 13.9% in hospital-based studies [24]. This discrepancy may be attributed to differences in study settings, smaller sample size, or under-detection due to limited screening practices observed during ANC visits. Indeed, less than half of the participants had their blood pressure measured during ANC, indicating potential missed opportunities for diagnosis.

Nutritional status findings indicate a dual burden of malnutrition, with 28% of women being underweight and 11.1% overweight. This reflects the ongoing nutritional transition in Bangladesh, where undernutrition and overnutrition coexist, particularly among vulnerable populations [25]. Similar patterns have been observed in other South Asian contexts, where maternal undernutrition remains high while overweight and obesity are increasing due to dietary and lifestyle changes [26]. Both extremes of BMI are associated with adverse pregnancy outcomes, emphasizing the need for routine nutritional assessment and counseling.

A critical finding of this study is the limited awareness of NCDs among pregnant women, with approximately two-thirds lacking knowledge about NCD-related risks and complications. This aligns with previous studies in Bangladesh that highlight low health literacy and limited awareness of chronic diseases among women, particularly in rural and disadvantaged communities [27]. Poor awareness may contribute to delayed care-seeking and underutilization of preventive services.

The study also identified substantial gaps in ANC service delivery related to NCD screening and counseling. Only a small proportion of women received information about NCDs, and none were counseled on risk factors or long-term implications. Furthermore, essential components such as blood glucose testing, family history assessment, and counseling on complications were inadequately performed. These findings are consistent with earlier reports indicating that maternal health programs in Bangladesh have historically focused on emergency obstetric care, with insufficient integration of NCD screening and management into routine ANC services [28].

The findings underscore the need to strengthen community-based and facility-based NCD screening within maternal health services. Integrating simple, cost-effective screening tools such as blood pressure measurement and OGTT at primary care level can facilitate early detection and management. Community health workers and midwives can play a crucial role in expanding access to these services, particularly in hard-to-reach populations such as teagarden communities. Similar integrated approaches have been recommended by global health authorities to address the growing burden of NCDs during pregnancy [23].

### Limitations

This study has several limitations. The small sample size and purposive sampling limit the generalizability of the findings. The facility-based design may introduce selection bias, as women not attending ANC services were not included. Additionally, the lack of analytical statistical methods limits the ability to explore associations between risk factors and outcomes.

### Implications for Practice and Policy

Despite these limitations, the study provides important insights into the burden of NCDs among a neglected population group. Strengthening ANC services to include routine NCD screening, improving awareness through community engagement, and ensuring follow-up care are essential steps. Policymakers should consider integrating NCD prevention and management into existing maternal health programs to achieve better maternal and neonatal outcomes.

## Conclusions

The major findings of the study include, 2% of the pregnant women in the teagardens were identified with high blood pressure, 11% were identified with overweight and 12.7% were found GDM positive (7.8 - <11 mmol/L). In ANC observation it was found only 4.2% cases receiving the NCDs related information where none of the cases found counseled about the NCD related risk factors. NCDs case could be identified with the adopted simple screening test which can be done by lower-level health workers. Other locally working NGOs may be engaged. Some of them are helping the community work; they can be used as formal partners at local level. Follow up of NCDs affected pregnant mothers after delivery need to strengthen and camping need to conduct periodically.

## Data Availability

All data produced in the present work are contained in the manuscript

